# Comparison of sedentary time calculated from count- and raw accelerometer data: The Tromsø Study

**DOI:** 10.1101/2025.06.02.25328840

**Authors:** Marc Weitz, Bente Morseth, Laila A. Hopstock, Søren Brage, Alexander Horsch

## Abstract

Accelerometers have become a popular and indispensable tool to objectively measure various aspects of daily behavior. While physical activity can be estimated relatively easy, sedentary behavior requires reliable Time in Bed and non-wear time algorithms. Over the years, various algorithms have been proposed, but little is known about how these algorithms quantitatively influence the estimation of sedentary behavior. In this study, we systematically compared different data processing strategies and their effect on sedentary behavior in 6155 participants asked to wear an accelerometer for seven consecutive days. We found considerable variation in the estimates ranging from 183 min to 807 min per day, mainly due to considerable influence by time-in-bed algorithms and sedentary thresholds. Non-wear time algorithms, on the other hand, only had a minor influence on the estimation of sedentary time, despite substantial variation between the algorithms. Our results suggest that sedentary behavior estimates from accelerometers must be interpreted with respect to the methods used to obtain them. When results from different studies are compared, the data should be reprocessed in a harmonized way.

## Introduction

Sedentary behavior commonly refers to a type of behavior during wake time spent in a seated or reclined posture. Sedentary behavior is also often characterized by little energy expenditure, commonly defined as less than 1.5 metabolic equivalent of task (MET) [1]. In fact, people from western cultures spend a considerable amount of their time sedentary [2, 3]. Whether health risks are associated with higher amounts of sedentary behavior or the lack of physical activity is an ongoing debate [4, 5]. Thus, it is an important prerequisite to accurately measure sedentary behavior and to distinguish sedentary time from other activities of light intensity.

A method that has gained great popularity to assess time-use behaviors like physical activity, sedentary behavior, and sleep in free-living settings is the use of accelerometers, also known as accelerometry or actigraphy [6]. Accelerometers are small, body-worn devices that measure raw acceleration (commonly expressed as multiples of Earth’s gravitation with 1 g = 9.806 65) along one to three axes. Modern accelerometers can record behavior over several days up to multiple weeks. In order to produce meaningful, interpretable results, the raw acceleration data needs to be aggregated and various algorithms need to be applied in order to identify the relevant recording times e.g. by excluding non-wear time. In the case of sedentary behavior, identification of sleep periods is also required as both behaviors produce comparable intensities, but are differentially associated with health and longevity [7, 8].

Thus, the first step in the processing of accelerometer data is the identification of non-wear time episodes. While some non-wear time is intended, e.g., the participants are instructed to remove the device during water contact (showering, swimming, etc.) or contact sports, often participants also remove the device in other situations and forget to put it on again. Many algorithms for count [9–11] and raw data [12–14] have been proposed and validated with mixed classification results [15, 16].

The identification of sleep episodes depends on the study protocol. While some studies instruct participants to remove the accelerometer overnight to intentionally skip sleep period identification, other studies employ protocols of continuous wear time with only few exceptions. Especially in these continuous wear protocols, which are now commonly used in epidemiological studies [17], the assessment of sedentary behavior might depend on the choice of the sleep identification algorithm. As of today, various sleep algorithms have been proposed, predominantly focusing on wrist attachment of the accelerometer. Only few have been developed or validated on hip-worn accelerometers [18]. Traditionally, the Cole-Kripke [19] and the Sadeh [20] algorithm have been used. Recently, we have presented an algorithm specifically for hip-worn accelerometers [18].

However, non-wear time and sleep algorithms have been commonly evaluated independently and it remains therefore unclear how combining sleep and non-wear time algorithms affects the estimation of sedentary time. For that reason, we apply different combinations of commonly used sleep/time in bed and non-wear time algorithms in this study and investigate the effect of method choice on the estimation of sedentary time. Additionally, we compare the estimated sedentary time between four demographic-specific and commonly used thresholds on three different metrics (uniaxial count, triaxial count and ENMO raw data).

## Method

### Participants

A total of 6155 accelerometer recordings was obtained from the seventh survey of the Tromsø Study [21]. The exact sampling procedure and the demographics of the data set have been described in detail elsewhere [22]. From the 6155 recordings, 52 raw data files could not be loaded or processed, leading to data of 6105 participants being included in this study. The original data collection had been approved by the Regional Committee for Medical Research Ethics (REC North ref. 2014/940) and the Norwegian Data Protection Authority, and all participants gave written informed consent. The usage of the Tromsø Study data has been approved by the Data and Publication Committee of the Tromsø Study (DPU 50/21) on 05.01.2021. The authors did not have access to information that could identify individual participants.

### Materials

The participants wore an ActiGraph wGT3X-BT (ActiGraph, Pensacola, FL), a small (4.6 × 3.3 × 1.5 cm) accelerometer, for eight consecutive days and nights on an elastic band on the right hip [21]. It was programmed to record raw acceleration in gravitational units g (1 g = 9.806 65 m s^−2^) at a sampling rate of 100 Hz with a dynamic range of ±8 g. Past the recording time, the participants returned the accelerometers by mail using prepaid envelopes.

### Procedure

Raw and count data was downloaded using ActiLife as GT3X and AGD files, respectively. The raw data was loaded via the Python package PAAT [23], while the count data could be loaded directly due to its CSV like structure. Accelerometer calibration correction coefficients were estimated using the R package GGIR [24].

These coefficients were used to autocalibrate the raw data using the method described by Van Hees et al. [25] for the metrics like ENMO [26]. We applied various non-wear time [9–14] and time in bed/sleep [18–20] algorithms. The count-based NWT algorithms were taken from the supporting repository of Syed et al. [15]. For the raw data based NWT algorithms [12, 14] and the time in bed algorithm of Weitz et al. [18] the implementations from PAAT were used. The count-based sleep algorithms [19, 20] were re-implemented based on the respective papers and can be found in the supporting repository. An overview of the used algorithms with brief descriptions is provided in Table 1. Physical activity levels were estimated by applying various demographic appropriate sedentary [27–30] and moderate-to-vigorous physical activity (MVPA) [27, 30–32] thresholds. The results from both, the raw and the count data, were combined into one file per person.

**Table 1.** Overview over the algorithms used in this study with the target variable and the metric the algorithm operates on. The algorithms and thresholds used in this study have been selected based on the demographics or previous application on this data.

| Method | Variable | Metric | Brief algorithm summary |
| --- | --- | --- | --- |
| Troiano et al. [9] | NWT | Uniaxial counts | Search for NWT intervals of at least 60 consecutive minutes of zero activity intensity counts, with allowance for 1–2 min of counts between 0 and 100 |
| Hecht et al. [10] | NWT | Triaxial counts | To be considered wear time, a the vector magnitude of one minute needs to fulfill at least two out of the following three constraints: (1) more than 5 CPM (2) at least 2 min with more than 5 CPM in the following 20 min (3) at least 2 min with more than 5 CPM in the preceding 20 min |
| Choi et al. [11] | NWT | Uniaxial counts | 1-min time intervals with consecutive zero counts for at least 90-min time window (window 1), allowing short time intervals with nonzero counts lasting up to 2 min (allowance interval) if no counts are detected during both the 30 min (window 2) of upstream and downstream from that interval; any nonzero counts except the allowed short interval are considered as wearing |
| Van Hees et al. [12, 13] | NWT | Triaxial raw | NWT based on standard deviation and value range of 30 min blocks of the accelerometer axes. The block is classified NWT if at least two out of the three axes have a standard deviation less than 3 mg or a value range of less than 50 mg |
| Syed et al. [14] | NWT | Triaxial raw | Convolutional neural network to detect when the accelerometer was removed and placed back on again |
| Cole et al. [19] | Sleep | Uniaxial counts | Sliding window approach over 7 min windows (four previous, the current, and two future minutes) where the counts of each minutes are weighted and summed up giving a score that is used to classify the current epoch |
| Sadeh et al. [20] | Sleep | Uniaxial counts | Calculates various metrics on a 11-minute window (current minute plus five previous and five future minutes) that are combined into one score indicating whether the the current minute is considered to be sleep |
| Weitz et al. [18] | TiB | Triaxial raw | A bidirectional long-short term memory network is used to classify raw 1 min acceleration time series into time in and out of bed |

ENMO: Euclidean Norm Minus One, NWT: Non Wear Time, TiB: Time in Bed, SB: Sedentary Behaviour, CPM: counts per minute

The 5 × 3 combinations of NWT and time in bed/sleep algorithms were used to exclude NWT and time in bed/sleep, respectively. For each combination the four different sedentary time thresholds were applied to the remaining data. For all 5 × 3 × 4 combinations, the periods labeled as sedentary time were summed up per day and participant. Subsequently, the data was further aggregated by taking the average of all days per participant and combination. The same approach was also applied to all NWT, TiB and sedentary time estimates independently to obtain how much the respective method would label when used in isolation.

## Results

Sedentary time estimates varied considerably between the different processing strategies (see Fig 1). The Hecht NWT algorithm together with the Sadeh sleep algorithm and the triaxial count data cutpoints [28] produced the lowest mean sedentary time estimate of 183.7 min (∼3 h, *SD* = 50.3), while the highest sedentary time estimate was obtained by using the Syed NWT algorithm together with the Weitz time in bed algorithm and the ENMO cutpoints proposed by Hildebrand et al. [29] estimating on average 806.9 min per day (∼ 13.5 h, *SD* = 121.6). A full overview over the sedentary time estimates is also provided in Table S3 Table.

**Fig 1.**
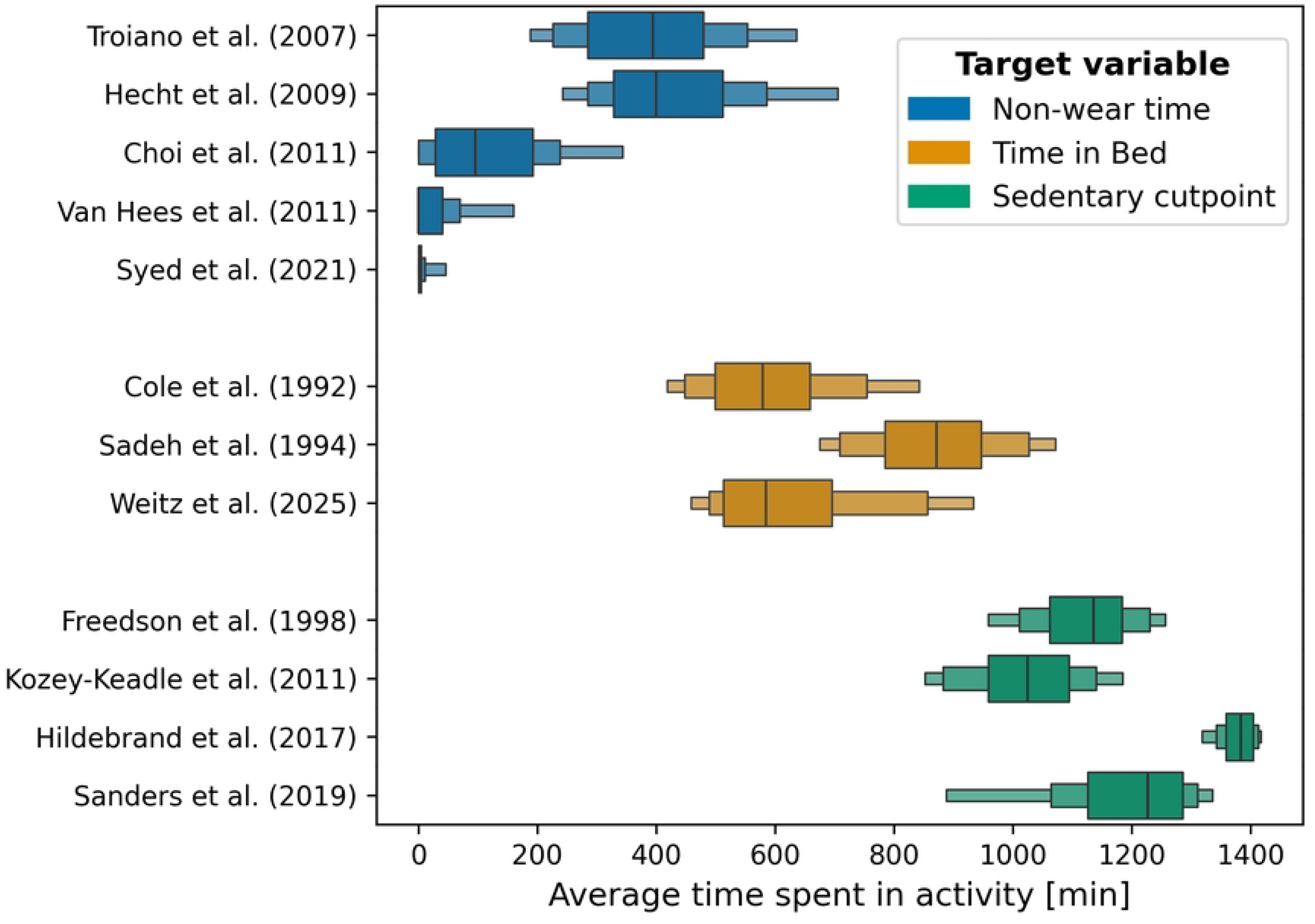
Letter-Value plots of the estimated sedentary time on the Tromsø Study data depending on the chosen NWT and TiB algorithm combination. Estimated sedentary time varied considerably ranging from 184 min to 807 min (3 h to 13.5 h) per day. The choice of NWT algorithm seemed to barely affect sedentary time estimation, while estimates seemed to be generally higher for the Cole-Kripke and Weitz TiB algorithm. The chosen cutpoint also seems to have a considerable influence with the Hildebrand cutpoint consistently estimating the highest volume of sedentary time. Data of 6105 participants is visualized with outliers being omitted for simplicity.

As shown in Fig 1, the choice of NWT algorithm seemed to barely influence sedentary time estimation, despite a huge variation of how much NWT was estimated (see Fig 2). TiB algorithms and the cutpoints, on the other hand, seemed to influence the sedentary time estimates considerably. It is worth noting, that the TiB algorithm of Weitz et al. [18] and the Cole-Kripke algorithm produced similar TiB estimates while the Sadeh algorithm estimated considerably more TiB on average per person (see Fig 2).

**Fig 2.**
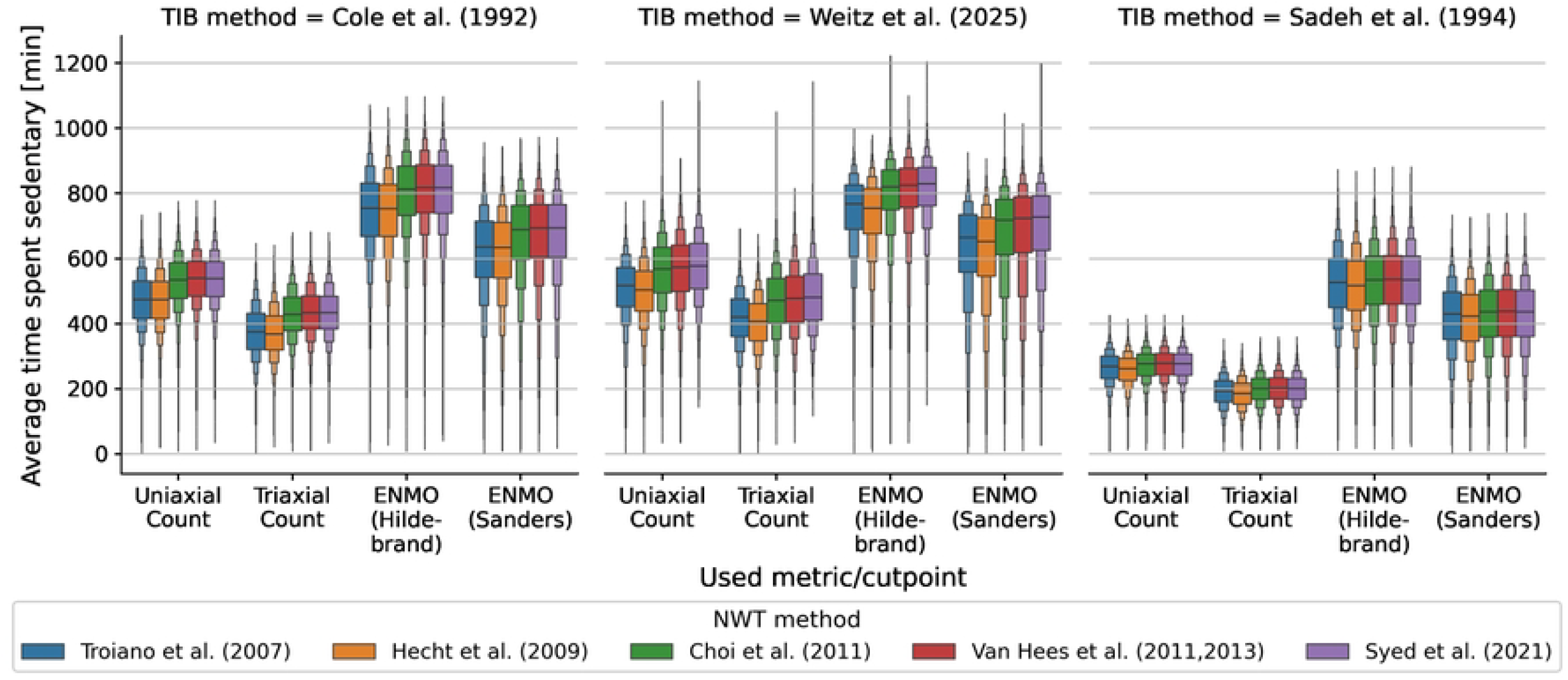
Average time spent in the respective activities estimated by the different used methods when applied to the data independently. For the NWT methods, early methods tend to estimate more NWT than more recent versions. For the TiB methods, the Cole-Kripke and the Weitz algorithm provided similar estimates on our data (600 min vs 622 min, respectively), while the Sadeh algorithm estimated considerably more TiB (∼ 876 min). For the cutpoints, triaxial count data produced the lowest sedentary time estimates, followed by uniaxial count data. The raw data based estimates seemed both to estimate higher volumes of daily sedentary behavior with the Hildebrand cutpoint labeling almost 96% as sedentary time. Data of 6105 participants is visualized with outliers being omitted for simplicity.

## Discussion

Our study demonstrated a considerable variation of estimated sedentary time between the different combinations of NWT algorithms, TiB algorithms and sedentary thresholds. The Sadeh algorithm labeled more time as TiB as the other two TiB algorithms, the Cole-Kripke and the Weitz algorithm, leading to lower volumes of sedentary behavior. It is worth noting, that despite the similar volumes of estimated TiB, a closer analysis revealed that the two methods label different parts of the data as TiB (see Fig S1 Fig).

The common goal of all investigated methods is to identify sedentary episodes during wake time defined by an energy expenditure ≤ 1.5 METs [1]. However, the considerable variation both in estimated volume but also in the periods labeled as sedentary behavior, indicates some shortcomings of the methodology. The differences can partially be explained by the different cutpoints used, a method which has been frequently criticized for its dependency on the population’s demographics and its limited comparability [33]. Even though all cutpoints used in this study have been selected to fit the demographics of the study population, or have been previously used on this data, the calibration protocols, procedures and samples between the used cutpoints still varied. An alternative to these calibration study based cutpoints, might be the usage of population based cutpoints as recently proposed by Migueles et al. [34].

A significant issue with comparing count and raw data based approaches is that they operate on different epoch durations. Cutpoints on count data have typically been developed for 1 min epochs [27, 28, 31]. Raw data based approaches often employ an epoch length of 1 s [29, 30, 32]. Epoch length, however, is known to influence physical activity estimation [35]. Orme et al. [35] showed that longer epoch length lead to lower non-bouted but higher bouted estimates of MVPA. While we could replicate this effect with our data, the results for sedentary time were more complex. While sedentary time estimates increased with longer epoch lengths for the Hildebrand cutpoint, they decreased for the Sanders cutpoint. One reason for this might be the significantly lower sedentary cutpoint proposed by Sanders et al. [30] (15 mg vs 47.4 mg). Nevertheless, these differences are still to small to explain the difference compared to the count data.

The advent of raw accelerometry led to many new methods published over the last decade. Commonly, these methods claimed to be more precise than previous methods given the richer and higher resolution data. Our results indicate considerable variation in sedentary time estimates depending on the selected non-wear time and time in bed algorithm. Here, we quantified the differences in commonly used algorithm combinations which facilitates comparisons between results from studies using different methodologies. It is worth noting in this context, that the investigated combinations also contain combinations of algorithms and cutpoints performed on different metrics, an approach that is uncommon in practice. However, even if taking only the combinations into account where algorithms and cutpoint were applied on the same metric, the general picture of considerable variation between sedentary time estimates remains.

It should therefore be of high importance to the field to interpret accelerometer-based sedentary time estimates with respect to the methods used to obtain them. If the goal is to compare two or multiple studies where raw data is available, harmonized analysis where all data is processed the same way, should be preferred. Where available, studies should also aim to publish the code used to process the data to facilitate harmonization and replication.

## Data Availability

The data of the Tromsø Study cannot be shared publicly to control for data sharing, including publication of datasets with the potential of reverse identification of de-identified sensitive participant information. Data are available from the Data and Publication Committee for the Tromsø Study (Contact information: The Tromsø Study, Department of Community Medicine, Faculty of Health Sciences, UiT The Arctic University of Norway) for researchers who meet the criteria for access of confidential data.

https://github.com/Trybnetic/method_comparison_paper

## Acknowledgements

This work was supported by the High North Population Studies at UiT The Arctic University of Norway. The publication charges for this article have been funded by a grant from the publication fund of UiT The Arctic University of Norway.

## Supporting information

**S1 Fig. Comparison between processing strategies reveals high variation of estimated activities.** (A) Sankey diagram visualizing the differences between triaxial count and raw data based processing on population level (*n* = 6105). (B) Visualization of the two processing strategies on one day data of one individual. Despite similar average TiB, NWT and MVPA estimates, both methods vary in which epochs are labeled respectively.

**S2 Fig. Influence of epoch length on estimated sedentary time and moderate-to-vigorous physical activity (MVPA).** Each point represents the average sedentary time of one of the non-wear time algorithm for the given cutpoint and time in bed algorithm. The bar indicate the average across the non-wear time algorithms. (A) Sedentary time estimates decreased with shorter epoch length for the Hildebrand cupoint, but increased for the Sanders cutpoint. (B) Estimated MVPA increased for both cutpoints with shorter epoch length.

**S3 Table. Estimated average sedentary time (and standard deviation) for all data processing strategies sorted by estimated volume.**

